# Reported harassment and mental-ill health in a Canadian prospective cohort of women and men in welding and electrical trades

**DOI:** 10.1101/2023.11.07.23298222

**Authors:** Jean-Michel Galarneau, Quentin Durand-Moreau, Nicola Cherry

**Affiliations:** Division of Preventive Medicine, University of Alberta, Edmonton, Canada; Faculty of Kinesiology, University of Calgary, Calgary, Canada

**Keywords:** Harassment, Workplace, Prospective studies, Anxiety, Depression, Welding

## Abstract

**Introduction:** Experience of psychosocial environments by workers entering trade apprenticeships may differ by gender. We aimed to document perceived harassment and to investigate whether this related to mental ill-health.

**Methods:** Cohorts of workers in welding and electrical trades were established, women recruited across Canada and men from Alberta. Participants were re-contacted every six months for up to 3 years (men) or 5 years (women). At each contact they were asked about symptoms of anxiety and depression made worse by work. After their last regular contact participants received a ‘wrap-up’ questionnaire that included questions on workplace harassment. In Alberta respondents who consented were linked to the administrative health database which recorded diagnostic codes for each physician contact.

**Results:** 1885 workers were recruited, 1001 in welding trades (447 women) and 884 in electrical trades (438 women). 1419 (75.3%) completed a ‘wrap up’ questionnaire, with 1413 answering questions on harassment. 60% of women and 32% of men reported that they had been harassed. Those who reported harassment had more frequently recorded episodes of anxiety and depression made worse by work in prospective data. In Alberta 1242 were successfully matched to administrative health records. Those who reported harassment were more likely to have a physician record of depression since starting in their trade.

**Conclusions:** Tradeswomen were much more likely than tradesmen to recall incidents of harassment. The results from record linkage, and from prospectively-collected reports of anxiety and depression made worse by work, support a conclusion that harassment resulted in poorer mental health.

**What is important about this paper?:** In this study we show that the majority of women entering the male-dominated trades of welding and electrical work report harassment, particularly during their apprenticeship, that is largely from co-workers and has a sexual component. Through its longitudinal design and linkage to an administrative health database, the study has reduced the impact of reporting bias and produced credible evidence that reported harassment is associated with anxiety and depression made worse by work and with physician reported depression. There is increasing recognition that workplace harassment of any worker is unacceptable, with obligations on the employer to take action to eliminate it. Evidence of ongoing mental health impacts reinforces this obligation.

## Introduction

Workplace harassment occurs worldwide and is explicitly addressed by the International Labor Organization Convention 190 [International Labor Organization, 2019] but its prevalence is not easily quantified, depending as it does on the operational definitions used and the types of behavior included [Cowie et al, 2002]. Questionnaires have been developed to study workplace violence [Leymann, 1996] and negative acts [Einarsen et al, 2009] or for wider psychosocial hazards, including bullying [Berthelsen et al 2020] but in practice there is little uniformity of approach, resulting in a wide range of prevalence estimates for national community-based samples. Using a set of questions unlikely to have elicited reports of sexual harassment, a prevalence of 17% was found for bullying in German workplaces, with rates higher in manual work and among younger employees: no difference in reported bullying was found between men and women [Lange et al,2019]. Sterud et al [2023] found a prevalence of 11% for all types of adverse social behavior (including sexual harassment) in Norway. In Australia a prevalence of 14.5% was found, in Australian-born workers, for bullying during the last year (9.9 % in men and 16.8% in women). The prevalence for ever having been bullied in the work place was 27% [Reid et al,2020]. In Hungary a 12-month prevalence of 49% was reported with higher rates in women and those aged 18-29 years [Szusecki et al, 2023]. In the United States a 12 month-prevalence of 60% for workplace violence was found in workers aged less than 25 years with higher rates in women (69%). Rates were lower in trades workers (23%) than in healthcare workers or customer service occupations [Rauscher et al,2023]. It is suggested that, in the United States, about 50% of women experience workplace sexual violence with increased risk in male dominated occupations [Riddle et al, 2023].

Few opportunities arise to compare employment experiences of women and men, closely matched on type of work. Studies have largely been conducted in large organizations with women in a minority (at least in certain roles) including among physicians [Frank et al, 1998] the military [Street et al, 2007] and firefighters [Jahnke et al,2019]. The WHAT-ME cohort [Cherry et al, 2018] comprised women and men recruited from welding and electrical trades. Although much of the study focus was on particulate and metal exposures [Galarneau et al, 2022], reproductive and other health outcomes [Cherry et al,2021] [Cherry et al, 2022], a final questionnaire included questions on workplace harassment. This paper describes the differing reports of harassment between women and men in these trades and also considers the relation between reported harassment and mental ill-health, acknowledging that interpretation of such data as causal is fraught with uncertainties. The relationship between mental ill-health and workplace psychosocial factors has been found to be strongest when both were measured during the same time period, raising questions about the direction of causality [Ahlin et al, 2019]. Meta-analyses suggest a positive relation between workplace bullying and depression [Verkuil et al, 2015] and other mental health disorders [Rudkjoebing et al, 2020], but with the same difficulty of interpretation. As with other self-reported workplace experiences [Kolstad et al, 2011], it seems likely that reporting bias inflates associations between bullying and depression. To reduce the likelihood of such bias, we considered the relation of reported harassment, collected retrospectively at the end of the study, to earlier reports of anxiety and depression made worse by work. We also used data linkage to assess the relation of self-reported harassment to physician reports of mental ill-health.

## Methods

### Cohort recruitment and follow-up

Four cohorts were set up, of women and men who had started an apprenticeship in a welding or electrical trade since 2005, as previously described [Cherry et al, 2018]. Recruitment of women began in January 2011 and of men in January 2014. To recruit sufficient women to address questions about reproductive outcomes, women were recruited from every Canadian province or territory, men just from Alberta. Participants were re-contacted every six months after recruitment for up to 3 years (men) or 5 years (women). Questionnaires were completed online or by telephone, in English or French. Along with questions on work, reproductive history and health, at each contact participants were asked if they were experiencing symptoms of asthma, rhinitis, dermatitis, shoulder pain, back pain, Raynaud’s phenomenon, anxiety or depression, and whether these were made worse by work (questions 1.1 to 1.8 supplementary materials 1). After a participant had completed their last regular contact they were sent a final ‘wrap-up’ questionnaire that included the Hospital Anxiety and Depression Scale (HADS) [Bjelland et al, 2002], questions on ethnicity (question 6.3 in supplementary materials 2) sexual orientation and gender identity (question 6.4), use of recreational drugs (questions 6.5-6.8) and workplace harassment (questions 4.1-4.43). A shorter version of the ‘wrap-up’ was used for those whose contact since baseline had been intermittent. No payment was made for participation in the study overall but participants were paid $20 Canadian for returning this final short form. The section on harassment was less structured for this group (question 5.1 in supplementary materials 3).

### Measurement of exposure to harassment

The full questionnaire asked separately about harassment during two time periods: the apprenticeship and during work in the trade as a certified craftsperson. Respondents were asked: *during your apprenticeship were you 1) ever subjected to psychological harassment; 2) ever subjected to physical violence; 3) ever subjected to sexual harassment?* with parallel questions for the post-apprenticeship period. Those who reported harassment were asked structured questions about each type of harassment and given open-text fields in which to provide details, if they wished. Inspection of the open-field responses suggested that the pre-coded options did not always accurately represent the events described and all responses, including those from the short questionnaire, were coded using a standard scheme, allowing for interpretation and incorporation of open-ended comments. The coding noted the work role of the harasser, the type of harassment and the response of the participant. Where it was unclear if an event occurred during or after apprenticeship, it was assigned to the apprenticeship period. Recoding of open-text fields was used only to reflect the details reported. All reports of harassment were retained as the exposure variable of interest.

### Measurement of mental ill-health

At each periodic contact, the respondent was asked: *do you have days when you feel sad, empty or depressed most of the time? Do you have days when you feel worried or anxious most of the time?* If the respondent said yes to either, they were asked: **Was this made worse by work?** These questions were added after the recruitment of women had started. They were present for all men but missing for one or more contacts for women recruited before April 2011. Such reports of symptoms are referred to as a ‘periodic anxiety report’ or ‘periodic depression report’. The analysis included, as the first key outcome, reports of anxiety or depression symptoms made worse by work.

In Alberta, information on mental ill-health prior to joining the trade and in the years since the start of trade work was available from linkage to the Alberta administrative health database for those who gave consent. In Alberta health care is free at the point of service. To be paid by the provincial health care insurance plan, a physician has to record at least one diagnosis. With ethical approval and participant consent these data can be made available for research. Physician reports for April 1^st^2002-March 31^st^2018 were extracted. Four variables were constructed i) any diagnosis of an anxiety, stress or adjustment disorder (ICD-9: 300.0 to 300.3, 300.5-300.9 308.0 to 308.9, 309.0 to 309.9: ICD-10: F40 to F43, F45, F48) before the date of joining the trade ii) any diagnosis of an anxiety, stress or adjustment disorder since the date of joining the trade iii) any diagnosis of a depressive disorder (ICD-9: 300.4 311; ICD-10: F32, F33. F34.1) before the date of joining the trade and iv) any diagnosis of a depressive disorder since the date of joining the trade.

The second key outcome for the Alberta sub-cohort was a physician diagnosis of an anxiety or depressive condition at any time since joining the trade.

### Measurement of confounders and effect modifiers

The HADS was completed during the long form of the wrap-up questionnaire. It includes 7 questions for anxiety and 7 for depression using a 4-point scale (0 to 3), with a score for each subscale ranging 0-21. The preamble asked respondents to consider how they had been feeling in the last week. In this analysis the scores obtained at the end of the study were used as a surrogate for scores that would have been obtained before entering the trade, had that been possible, and so to adjust for a personal characteristic that might relate to reporting harassment. Such an approach is conservative in that it does not allow for changes to HADS scores that might have arisen through harassment.

Other potential confounders were trade (welding or electrical) and gender (woman or man) recorded by the apprenticeship board. For the Alberta sub-cohort, a physician diagnosis of an anxiety or depressive condition at any time before joining the trade was included as a potential effect modifier.

### Statistical methods

Prevalence of reporting any harassment since starting in the trade was calculated overall and for women and men by trade, by period of employment (apprenticeship or as a certified craftsperson). Episodes reported as harassment were examined by the work role of the harasser (supervisor or coworker) and by whether or not the incident included a sexual component. Effects on mental health were examined by the proportion of questionnaires on which the respondent recorded having symptoms of periodic anxiety or periodic depression made worse be work. This was calculated as a percentage of the total responses for that participant. As participants had periods not working or working in other jobs, computation of episodes made worse by work included only those in their trade at the time of reporting. A probit (fractional) regression examined factors associated with the proportion of periodic reports of anxiety or depression made worse by work. Multilevel multivariable logistic regression was used to assess the relation of reported harassment to episodes with anxiety or depression made worse by work. Differences by gender were explored by stratification. Further logistic regression analyses, using data from the administrative health database, took as the outcome variables physician-recorded anxiety or depressive disorders, with reported harassment as the exposure of interest and adjusting for the same disorder recorded in the health database before joining the trade. The analyses were carried out in Stata (StataCorp. 2015. *Stata Statistical Software: Release 14*. College Station, TX: StataCorp LP).

## Results

1885 tradespeople completed the baseline (recruitment) questionnaire, 1001 in welding trades and 884 in electrical trades. 447 in welding were women with 438 in the electrical trades. Mean age at starting apprenticeship was 25.5 years. In the whole cohort 75.0% (1413/1885) completed the harassment section on either the long (976) or short (437) follow-up questionnaire, including 691 women. (Figure 1) There were 1252 in the Alberta sub-cohort data-linked with the administrative health database: 946/1252 (75.6%) answered the harassment questions (Figure 2). Almost all (97%) of those completing the harassment questions on the long questionnaire also completed the HADS questionnaire (948/976 in the whole cohort 665/686 in the Alberta sub-cohort). Of those completing the long questionnaire, 45 identified as Indigenous People, 43 identified as LGBTQ+ and 88 reported using recreational drugs at least weekly.

**Figure 1.**
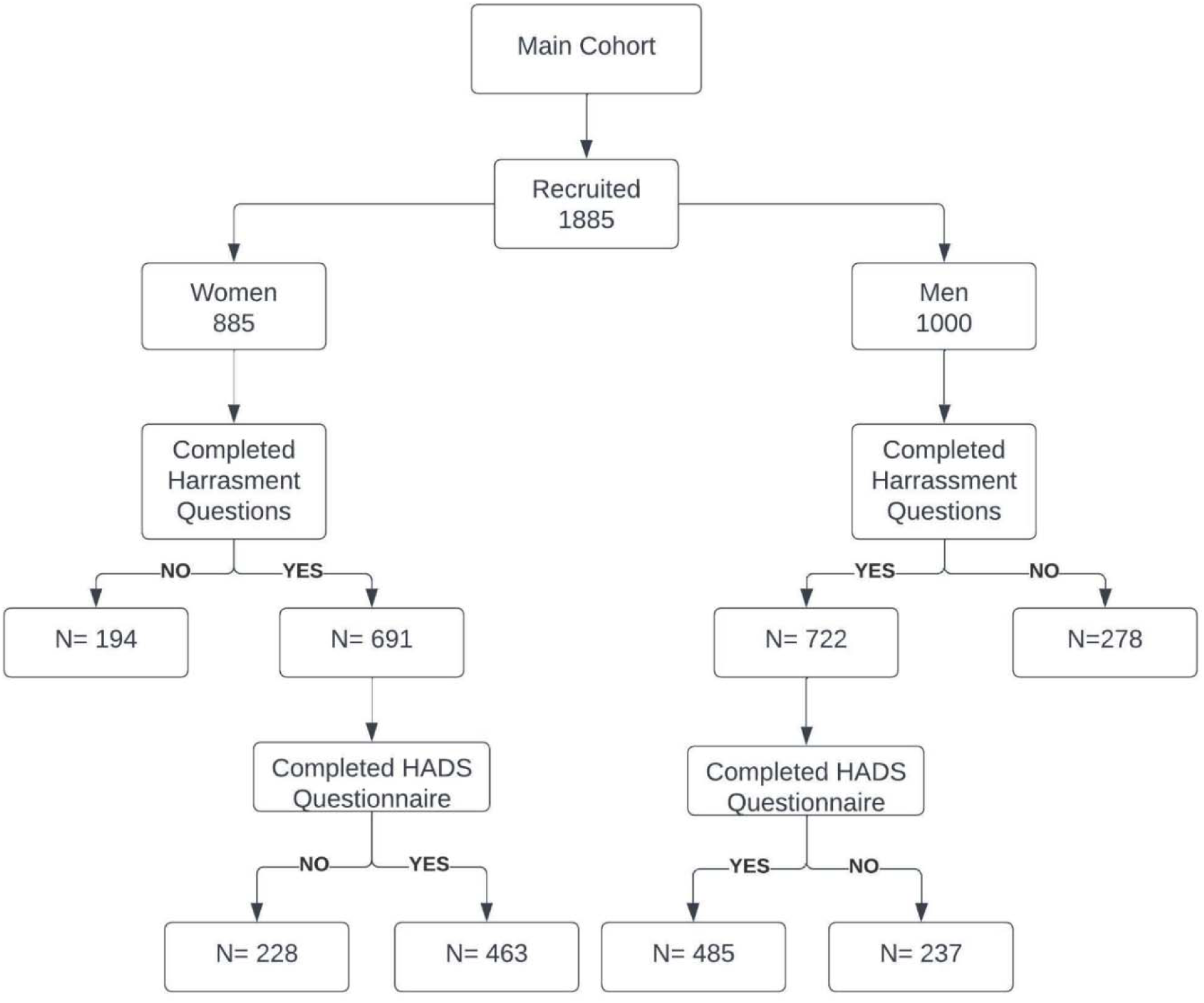
Flowchart for the main study cohort

**Figure 2.**
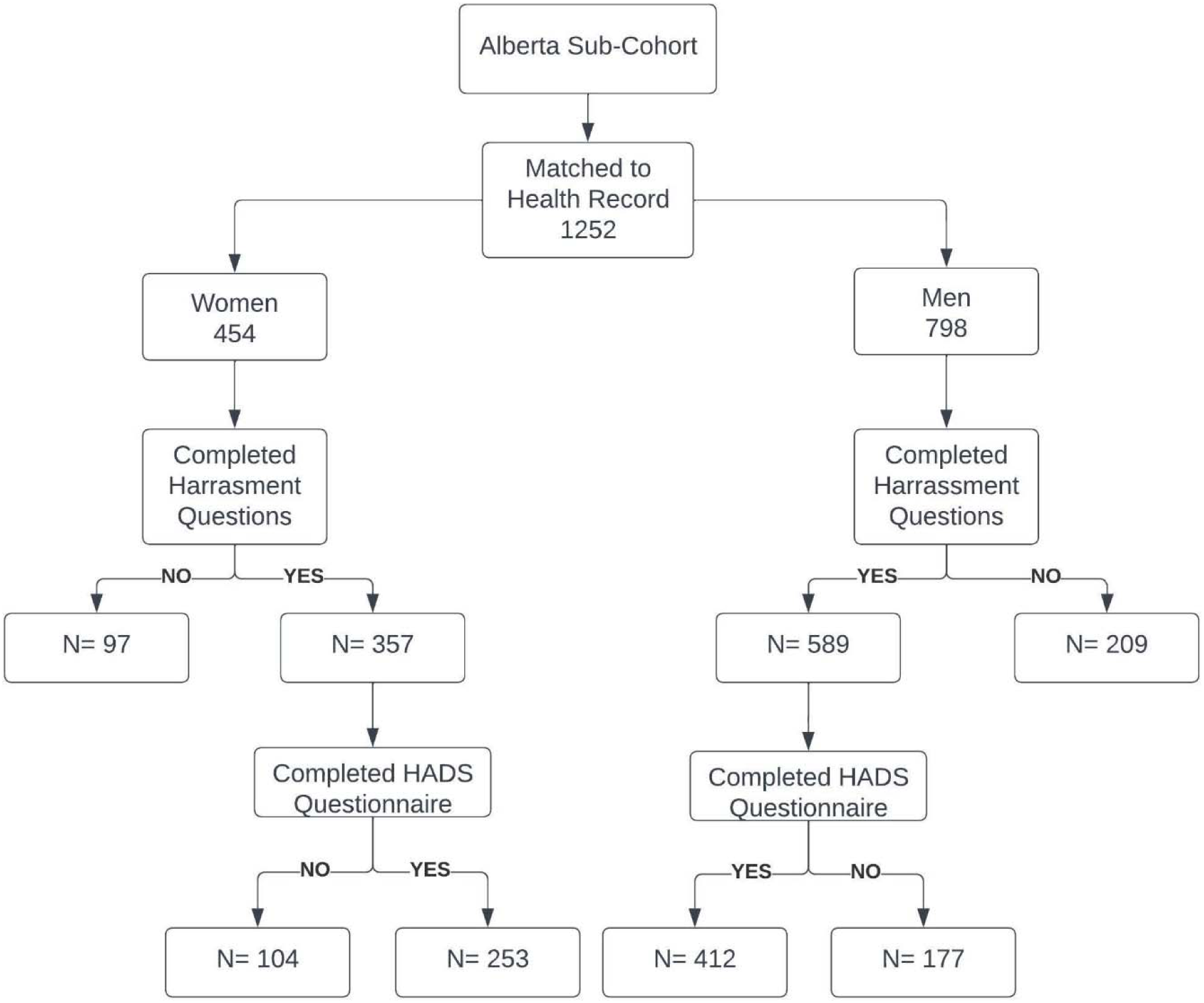
Flowchart for the Alberta sub-cohort

Overall (Table 1) 45.3% reported that at some time since starting their trade they had experienced harassment. The proportions were similar for welding (47.1%) and electrical trades (43.5%) but markedly higher for women (59.5%) than men (31.7%). Among those completing the long ‘wrap up’, the proportions reporting harassment during apprenticeship were similar to the overall estimate (62.1% in women, 32.3% in men) but lower for the period ‘in trade’ (48.3% in women, 22.9% in men). Those identifying as Indigenous People reported somewhat greater harassment overall (62.2%; Fishers exact test: p=0.126) as did those identifying as LGBTQ+ (69.8%; Fishers exact test p=0.012) or being frequent users of recreational drugs (61.4%; Fishers exact test: p=0.033). More women than men reported being harassed by their supervisor and, particularly, by coworkers (50% of women and 22% of men during their apprenticeship). Other harassers were mentioned much less frequently. Much of the reported harassment had a sexual component with 44% of women reporting this in relation to their apprenticeship and 28% once qualified and working in the trades (Table 1).

**Table 1:** Report of harassment by gender, trade, type of harassment and employment period.

| Harassment period | All |  |  | Women |  |  | Men |  |  |
| --- | --- | --- | --- | --- | --- | --- | --- | --- | --- |
|  | N | Harassed = yes | % | N | Harassed = yes | % | N | Harassed = yes | % |
| Any* |  |  |  |  |  |  |  |  |  |
| Welding | 709 | 334 | 47.1 | 336 | 205 | 61.0 | 373 | 129 | 34.6 |
| Electrical | 704 | 306 | 43.5 | 355 | 206 | 58.0 | 349 | 100 | 28.7 |
| Both | 1413 | 640 | 45.3 | 691 | 411 | 59.5 | 722 | 229 | 31.7 |
| During apprenticeship** |  |  |  |  |  |  |  |  |  |
| Welding | 454 | 210 | 46.3 | 206 | 128 | 62.1 | 248 | 82 | 33.1 |
| Electrical | 520 | 246 | 47.3 | 269 | 167 | 62.1 | 251 | 79 | 31.5 |
| Both | 974 | 456 | 46.8 | 475 | 295 | 62.1 | 499 | 161 | 32.3 |
| Who? |  |  |  |  |  |  |  |  |  |
| Supervisor | 974 | 265 | 27.2 | 475 | 157 | 33.3 | 499 | 108 | 21.6 |
| Co-worker | 974 | 348 | 35.7 | 475 | 237 | 50.2 | 499 | 111 | 22.2 |
| How? |  |  |  |  |  |  |  |  |  |
| Sexual | 974 | 224 | 23.0 | 475 | 208 | 43.8 | 499 | 16 | 3.2 |
| In trade, post-apprenticeship** |  |  |  |  |  |  |  |  |  |
| Welding | 359 | 136 | 37.9 | 156 | 82 | 52.6 | 203 | 54 | 26.6 |
| Electrical | 433 | 140 | 32.3 | 217 | 98 | 45.2 | 216 | 42 | 19.4 |
| Both | 792 | 276 | 34.8 | 373 | 180 | 48.3 | 419 | 96 | 22.9 |
| Who? |  |  |  |  |  |  |  |  |  |
| Supervisor | 792 | 146 | 18.4 | 373 | 82 | 22.0 | 419 | 64 | 15.3 |
| Co-worker | 792 | 196 | 24.7 | 373 | 139 | 37.3 | 419 | 57 | 13.6 |
| How? |  |  |  |  |  |  |  |  |  |
| Sexual | 792 | 109 | 13.8 | 373 | 105 | 28.2 | 419 | 4 | 1.0 |

\*Those completing the long or short harassment questions.
\*\*Those completing the long harassment questions only.

Among those who had completed the HADS questionnaire as part of the final contact, participants reporting harassment had higher scores on both the anxiety and depression scales, with similar patterns for men and women (top panel of Table 2).

**Table 2:** Anxiety and depression from the Hospital Anxiety and Depression Scale (HADS) and proportion of periodic reports by report of anxiety or depression made worse by work by harassment reported on ‘wrap-up’ questionnaire.

| Any report of harassment | HADS Score |  |  |  |  |  |  |
| --- | --- | --- | --- | --- | --- | --- | --- |
|  | N | Anxiety |  |  | Depression |  |  |
|  |  | Median | Mean | SD | Median | Mean | SD |
| Women |  |  |  |  |  |  |  |
| No | 163 | 5.0 | 5.5 | 3.4 | 2.0 | 3.0 | 2.6 |
| Yes | 300 | 8.0 | 7.8 | 4.3 | 4.0 | 4.4 | 3.5 |
| All | 463 | 7.0 | 7.0 | 4.2 | 3.0 | 3.9 | 3.2 |
| Men |  |  |  |  |  |  |  |
| No | 313 | 5.0 | 5.1 | 3.4 | 3.0 | 3.5 | 3.1 |
| Yes | 172 | 7.0 | 7.0 | 3.9 | 5.0 | 5.4 | 4.1 |
| All | 485 | 5.0 | 5.8 | 3.7 | 3.0 | 4.2 | 3.6 |
| Overall |  |  |  |  |  |  |  |
| No | 476 | 5.0 | 5.2 | 3.4 | 2.0 | 3.3 | 3.0 |
| Yes | 472 | 7.0 | 7.5 | 4.2 | 4.0 | 4.7 | 3.7 |
| All | 948 | 6.0 | 6.4 | 4.0 | 3.0 | 4.0 | 3.4 |
| Report of harassment | All with periodic reports while in trade |  |  |  |  |  |  |
|  | N* | Proportion of reports with anxiety made worse by work % |  |  | Proportion of reports with depression made worse by work % |  |  |
|  |  | Median | Mean | SD | Median | Mean | SD |
| Women |  |  |  |  |  |  |  |
| No | 131 | 0.0 | 7.3 | 15.7 | 0.0 | 3.8 | 13.3 |
| Yes | 246 | 0.0 | 21.6 | 31.8 | 0.0 | 15.3 | 27.4 |
| All | 377 | 0.0 | 16.7 | 28.1 | 0.0 | 11.3 | 24.1 |
| Men |  |  |  |  |  |  |  |
| No | 288 | 0.0 | 10.5 | 20.8 | 0.0 | 7.7 | 18.4 |
| Yes | 157 | 14.3 | 24.7 | 30.2 | 0.0 | 18.7 | 29.4 |
| All | 445 | 0.0 | 15.5 | 25.4 | 0.0 | 11.6 | 23.5 |
| Overall |  |  |  |  |  |  |  |
| No | 419 | 0.0 | 9.5 | 19.4 | 0.0 | 6.5 | 17.1 |
| Yes | 403 | 0.0 | 22.8 | 31.2 | 0.0 | 16.6 | 28.2 |
| All | 822 | 0.0 | 16.0 | 26.7 | 0.0 | 11.5 | 23.8 |

Note: All HADS scores for those reporting and not reporting harassment differed with $p < 0.001$
\*Participants completing the HADS questionnaire and periodic reports while in trade

The average numbers of questionnaires with a response to questions about periodic anxiety and depression were 5.2 (median 6) for men and 4.6 (median 4) for women, reflecting the late inclusion of the question for women and their greater likelihood of leaving the trade or taking time away for childbirth or other reasons. At the time of the ‘wrap-up’ questionnaire 25% (182/722) of men but 43% (296/691) of women reported not currently working in their trade. While in their trade 37.8% (168/445) of men reported at least once that they had anxiety made worse by work and 28.3% (126/445) depression made worse by work. The proportions for women were similar, 35.5% (134/377) and 25.2% (95/377). Those reporting harassment on the ‘wrap-up’ questionnaire had a higher proportion of periodic episodes made worse by work. This was seen for both women and men (lower panel of Table 2). In a fractional regression analysis, the proportions made worse by work were significantly higher for those reporting harassment (Table 1 in supplementary materials 4).

The higher proportion with symptoms made worse by work could have arisen either from reporting more anxiety or depression, without increased work attribution, or from the same levels of anxiety or depression but with a greater attribution to work. Table 3 considers first the relationship between periodic anxiety and depression and reported harassment. The odds ratio for harassment was always greater than 1.00 but not significantly so for anxiety in women. Type of trade did not contribute (except marginally for depression in women welders). Men were more likely than women to have periodic reports of anxiety, having adjusted for HADS anxiety score. The lower part of Table 3 gives the odds that a reported episode of anxiety or depression would be attributed to work. Here reported harassment was strongly related to work attribution of both anxiety and depression overall and in women. For men, while the odds ratios were clearly above 1 (1.67 for anxiety and 1.53 for depression) these had a probability >0.05. The analyses in Table 3 were then repeated for the Alberta subsample with adjustment for mental ill-health prior to joining their trade, taken from the administrative health database. Among the 1252 matched, 16.6% (208/1252) had an anxiety related condition before entering the trade and 13.3% (167/1252) a depressive condition (Table 2 supplementary materials 4). No relation was found between previous mental ill-health and the likelihood of reporting anxiety or depression, or such a condition made worse by work. (Table 3, supplementary materials 4).

**Table 3:** Multilevel logistic regression of periodic reports of anxiety and depression while in trade.

|  | Periodic reports of anxiety or depression while in trade |  |  |  |  |  |  |  |  |
| --- | --- | --- | --- | --- | --- | --- | --- | --- | --- |
|  | All |  |  | Women |  |  | Men |  |  |
| Anxiety | OR | 95% CI | P= | OR | 95% CI | P= | OR | 95% CI | P= |
| Harassment reported | 1.86 | 1.28 to 2.71 | <0.001 | 1.18 | 0.68 to 2.05 | 0.567 | 2.55 | 1.56 to 4.16 | <0.001 |
| HADS anxiety | 1.37 | 1.30 to 1.44 | <0.001 | 1.35 | 1.26 to 1.45 | <0.001 | 1.39 | 1.30 to 1.49 | <0.001 |
| Trade: welding | 0.95 | 0.67 to 1.36 | 0.797 | 0.88 | 0.53 to 1.46 | 0.617 | 1.00 | 0.61 to 1.63 | 0.988 |
| Gender: male | 1.55 | 1.07 to 2.23 | 0.020 | - | - | - | - | - | - |
| Depression |  |  |  |  |  |  |  |  |  |
| Harassment reported | 2.79 | 1.75 to 4.45 | <0.001 | 4.35 | 2.11 to 8.96 | <0.001 | 2.24 | 1.22 to 4.12 | 0.009 |
| HADS depression | 1.37 | 1.29 to 1.45 | <0.001 | 1.42 | 1.29 to 1.55 | <0.001 | 1.38 | 1.27 to 1.49 | <0.001 |
| Trade: welding | 1.43 | 0.95 to 2.15 | 0.086 | 1.65 | 0.95 to 2.85 | 0.074 | 1.30 | 0.74 to 2.29 | 0.364 |
| Gender: man | 1.06 | 0.71 to 1.58 | 0.786 | - | - | - | - | - | - |
| N observations | 4027 | - | - | 1718 | - | - | 2309 | - | - |
| Participants | 822 | - | - | 377 | - | - | 445 | - | - |
|  | Periodic reports of anxiety or depression made worse by work (in those reporting symptoms) |  |  |  |  |  |  |  |  |
|  | All |  |  | Women |  |  | Men |  |  |
| Anxiety | OR | 95% CI | P= | OR | 95% CI | P= | OR | 95% CI | P= |
| Harassment reported | 2.04 | 1.27 to 3.29 | 0.003 | 2.64 | 1.27 to 5.48 | 0.009 | 1.67 | 0.88 to 3.15 | 0.115 |
| HADS anxiety | 1.14 | 1.07 to 1.21 | <0.001 | 1.16 | 1.07 to 1.26 | 0.001 | 1.11 | 1.01 to 1.21 | 0.028 |
| Trade: welding | 0.69 | 0.44 to 1.08 | 0.104 | 0.69 | 0.36 to 1.32 | 0.258 | 0.68 | 0.36 to 1.27 | 0.226 |
| Gender: man | 2.01 | 1.25 to 3.23 | 0.004 | - | - | - | - | - | - |
| N observations | 944 | - | - | 420 | - | - | 524 | - | - |
| Participants | 397 | - | - | 184 | - | - | 213 | - | - |
| Depression |  |  |  |  |  |  |  |  |  |
| Harassment reported | 2.31 | 1.30 to 4.10 | 0.004 | 4.97 | 2.04 to 12.09 | <0.001 | 1.53 | 0.73 to 3.19 | 0.259 |
| HADS depression | 1.09 | 1.01 to 1.17 | 0.023 | 1.09 | 0.98 to 1.22 | 0.120 | 1.09 | 0.99 to 1.19 | 0.077 |
| Trade: welding | 0.97 | 0.58 to 1.64 | 0.920 | 1.21 | 0.59 to 2.48 | 0.597 | 0.85 | 0.41 to 1.75 | 0.652 |
| Gender: male | 1.45 | 0.83 to 2.55 | 0.194 | - | - | - | - | - | - |
| N observations | 723 | - | - | 312 | - | - | 411 | - | - |
| Participants | 324 | - | - | 143 | - | - | 181 | - | - |

An additional analysis considered whether the type of harassment (sexual or other) was associated with differences in reports of periodic anxiety or depression and their attribution to work. For women differences in odds ratios between sexual or other harassment were small but much larger increases were found, for both anxiety and depression, for the small numbers of men reporting sexual harassment (Table 4, upper panel, supplementary materials 4) although this increase among men reporting sexual harassment was not reflected in their attribution to work (Table 4 lower panel, supplementary materials 4).

**Table 4:**
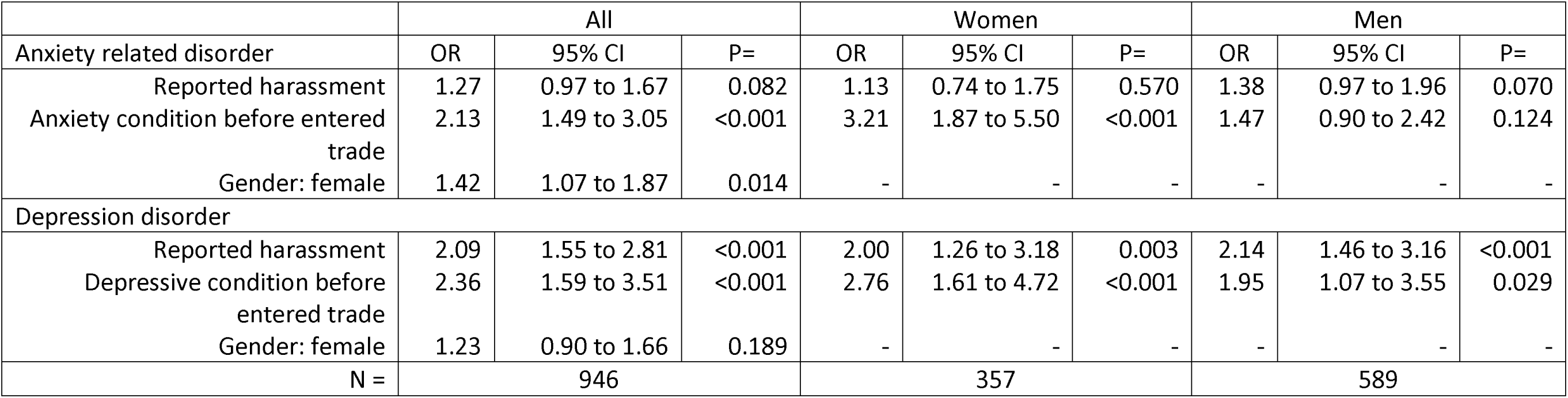
Relationship of harassment to physician diagnosed depression and anxiety disorder since joining the trade (logistic regression).

|  | All |  |  | Women |  |  | Men |  |  |
| --- | --- | --- | --- | --- | --- | --- | --- | --- | --- |
| Anxiety related disorder | OR | 95% CI | P= | OR | 95% CI | P= | OR | 95% CI | P= |
| Reported harassment | 1.27 | 0.97 to 1.67 | 0.082 | 1.13 | 0.74 to 1.75 | 0.570 | 1.38 | 0.97 to 1.96 | 0.070 |
| Anxiety condition before entered trade | 2.13 | 1.49 to 3.05 | <0.001 | 3.21 | 1.87 to 5.50 | <0.001 | 1.47 | 0.90 to 2.42 | 0.124 |
| Gender: female | 1.42 | 1.07 to 1.87 | 0.014 | - | - | - | - | - | - |
| Depression disorder |  |  |  |  |  |  |  |  |  |
| Reported harassment | 2.09 | 1.55 to 2.81 | <0.001 | 2.00 | 1.26 to 3.18 | 0.003 | 2.14 | 1.46 to 3.16 | <0.001 |
| Depressive condition before entered trade | 2.36 | 1.59 to 3.51 | <0.001 | 2.76 | 1.61 to 4.72 | <0.001 | 1.95 | 1.07 to 3.55 | 0.029 |
| Gender: female | 1.23 | 0.90 to 1.66 | 0.189 | - | - | - | - | - | - |
| N = | 946 |  |  | 357 |  |  | 589 |  |  |

The final table (Table 4) reports on the relationship between reported harassment and diagnoses recorded in the administrative health database. The outcome variables were diagnoses since joining the trade. Among 946/1252 who had completed the harassment questions, 47.6% had been recorded by a physician as having an anxiety-related condition since joining their trade and 29.1% a depressive condition (Table 5 supplementary materials 4). From Table 4, reported harassment was related to physician-diagnosed depressive disorders in both men and women but the relationship to physician-diagnosed anxiety disorders was much weaker.

## Discussion

This report had three objectives. The first was to describe and compare the reported experiences of workplace harassment in women and men, closely matched on type of work, in trades traditionally entered by men. It found substantial reports of harassment in both genders, particularly during apprenticeship training. Levels were markedly higher in women, with harassment by coworkers, and behavior with a sexual component being particularly in evidence. The second objective was to use the data collected prospectively to contribute to discussion of the effects of harassment on mental ill-health. Although missing some data that would have helped interpretation, the observed relationship between post-hoc reporting of harassment and the periodic reports, collected prospectively, of anxiety and depression made worse by work, cannot easily be attributed solely to bias or unmeasured confounding. Third, for those linked to the Alberta administrative health database, the analysis assessed the contribution of harassment to physician recorded mental ill-health, so removing the element of health self-report. The results support the contribution of reported harassment, particularly to depression, in both women and men.

The strengths of the study include the recruitment and retention of a large cohort of women and men going through the same apprenticeship training in the welding or electrical trades and, largely, working in the same trades post-apprenticeship. A further strength is the repeated prospective collection of symptoms of anxiety and depression and reports of the role of work in making those symptoms worse. The link to administrative health data is an additional strength.

There also limitations, both for the study overall and for its ability to answer the questions central to this analysis. First, as discussed elsewhere [Cherry et al, 2018], the response to the invitation to take part in the study was low (5 −15%) and those taking part may differ importantly from the whole cohort starting an apprenticeship since 2005. Second, although a 75% response rate to the final questionnaire is high for a follow-up study covering several years [Graaf et al, 2013] [Njuyen et al, 2023], inclusion in the central analyses reported here was limited to those 948 completing the HADS scale (50% of those recruited). Similarly, information on ethnicity and gender identification was collected only in the long wrap-up and small numbers limit discussion of the outcomes of harassment in these subgroups (although this might be expected [Durand-Moreau et al 2022). Moreover, the information on gender, derived from apprenticeship records, did not allow consideration of a gender spectrum. A further limitation is the absence of data on when the harassment occurred: we could not link harassment events to a specific report of anxiety or depression made worse by work or to a date of physician diagnosis, nor can we estimate 12-month prevalence rates to compare with other studies. Data on mental health before the study wer limited to administrative health reports in the Alberta cohort. Although the use of physician records is a strength, there are limitations to these data. Physicians record billing codes that correspond to their clinical impression and it cannot be assumed they carried out a full mental health assessment. Further the physician database would not include mental health assessments by a psychologist or counsellor.

The inherent difficulty in studies of the effects of harassment on mental health lies in the measurement of harassment. While it may be possible to get independent or objective measures of mental health (as here), use of outside perspectives [Cowie et al, 2002] [Gullander et al, 2014] to record harassment has important limitations, particularly perhaps for sexual harassment and other behaviors not in the public view. In the present study, involving hundreds of different workplaces, measurement other than self-report was not feasible. Self-report is a strength in that the person experiencing workplace events as harassing is presumptively the best source of information about the event (with external reality checks, if available), but reliance on self-report has particular difficulties for assessing the causal link (if any) between harassment and later mental ill-health. Both experience and recollection of harassment may reflect current mental health. Someone who is anxious or depressed for reasons unconnected to work may find workplace frictions to be harassing to a degree they would not have experienced without the external stressors. Similarly, recollection of workplace events may be colored by current mental state, with those now anxious or depressed being more likely to interpret past frictions in a negative light. Further, a report of harassment may reflect dissatisfactions about working conditions more generally, rather than specific harassing behaviors. The design of the present study counters most of these potential biases.

The collection of self-report data on harassment only at the end of the study avoided biases that would have arisen from the simultaneous recording of exposure and effect. The possibility of a long-standing susceptibility to mental-ill-health, affecting both workplace anxiety/depression and perception of harassment, has been addressed by adjustment for HADS anxiety and depression scores. It remains possible that a negative relationship to work may have influenced both the prospective reporting (from 2011) of anxiety or depression as work related and the retrospective report of harassment (from 2016, after completion of the last prospective report). A simplified analysis (supplementary materials 5) suggests that it would have needed 38% of false positives (106/278 reports of harassment that did not occur) among those who had reported either anxiety or depression made worse by work for such a bias to negate the findings reported here The term ‘harassment’ used here does not necessarily conform to definitions used by other researchers or by governmental bodies, considering the possibility of enforcement. Harassment is defined for Canadian federal employees as “any action, conduct or comment, including of a sexual nature, that can reasonably be expected to cause offence, humiliation or other physical or psychological injury or illness to an employee” [Government of Canada, 2020]: we do not know if, in a situation of enforcement, the harassment reported here could reasonably be expected to cause offence. Other definitions require that actions occur repeatedly over a longer period [Einarsen et al,2009]. Since we did not ask this we cannot quantify the proportion in which the actions occurred repeatedly, although the comments recorded in the open-text field suggest this was high. Moreover, self-reported harassment may in part reflect other issues at the workplace, unmeasured in this study [Durand-Moreau and Lasfargues.2022].

The analysis reported here suggests that events at work (presumptively including harassment) can worsen mental health, perhaps only transiently, but resulting in physician diagnoses of depressive conditions. Given the high rates of reported harassment in women in these trades traditionally entered by men (well above working lifetime prevalence in women in Australia, for example [Reid et al, 2020] but comparable to those reported in Hungary [Szusecki et al 2023]and the United States [Rauscher et al, 2023] [Riddle et al, 2023]), the goal must be to change cultures such that harassment becomes unacceptable. A Cochrane review [Gillen et al, 2017] of existing interventions to prevent bullying was not encouraging but the experiences reported by these women and men are clearly unacceptable in any society and are certainly contrary to the wording and intent of provincial [Government of Alberta Occupational Health and Safety, 2023] and federal [Government of Canada, 220] legislation and the International Labor Organization convention [2019].

## Ethics

The project was reviewed by the Health Ethics Review Board of the University of Alberta (Pro00017851). All participants gave written informed consent.

## Data availability statement

The data underlying this article will be shared on reasonable request to the corresponding author.

## Contributor statement

NC devised the study and lead the research team. J-M G created the data bases used in the analysis. Q D-M was integral to the conception and coding of the details of reported harassment. All contributed to the analysis and writing of the report.

## Acknowledgements

Laura Rodgers played a major role in contacting the participants over the several years of follow-up and in obtaining participation in the ‘wrap-up questionnaire.

