## Supplementary materials for "Reported harassment and mental-ill health in a Canadian prospective cohort of women and men in welding and electrical trades"

Page

1) Health questions from the follow-up questionnaire for those in electrical trades	2
2) Long wrap-up questionnaire	6
3) Short wrap-up questionnaire	20
4) Tables SM1-SM5	23
5) Calculation of misreporting needed to eliminate a result. 29

Reported harassment and mental-ill health in a Canadian prospective cohort of women and men in welding and electrical trades

Jean-Michel Galarneau, Quentin Durand-Moreau, Nicola Cherry

**Supplementary materials 1**

Health questions from the follow-up questionnaire for those in electrical trades

**Electricians Follow-Up Questionnaire**

**Thank you for taking the time to fill in this questionnaire. Could I first ask some questions about how your health is now?**

1 **Do you have any of the following health problems now?**

| Do you have: |  |  | How much have you been  troubled by this in the last month? Score this on a scale of 1 to 10 where 1 means troubled very little and 10 means extremely troubled. | Was this made worse by work? | |
| --- | --- | --- | --- | --- | --- |
| 1.1 Asthma or wheezing when you ***do not*** have a cold? | Yes  No | **□** 1  **□** 2 |  | Yes  No | **□** 1  **□** 2 |
| 1.2 Sneezing, a runny, or a blocked nose when you ***do not*** have a cold ? | Yes  No | **□** 1  **□** 2 |  | Yes  No | **□** 1  **□** 2 |
| 1.3 Dermatitis or  itchy/inflamed skin when you ***do not*** have insect bites or sunburn? | Yes  No | **□** 1  **□** 2 |  | Yes  No | **□** 1  **□** 2 |
| 1.4 Shoulder pain sufficient that it interferes with your normal activities | Yes  No | **□** 1  **□** 2 |  | Yes  No | **□** 1  **□** 2 |
| 1.5 Low back pain sufficient that it interferes with your normal activities | Yes  No | **□** 1  **□** 2 |  | Yes  No | **□** 1  **□** 2 |
| 1.6 Episodes when your fingers turn white, with numbness or tingling, in the cold? | Yes  No | **□** 1  **□** 2 |  | Yes  No | **□** 1  **□** 2 |
| 1.7 Days when you feel sad, empty, or depressed most of the time? | Yes  No | **□** 1  **□** 2 |  | Yes  No | **□** 1  **□** 2 |
| 1.8 Days when you feel worried or anxious most of the time? | Yes  No | **□** 1  **□** 2 |  | Yes  No | **□** 1  **□** 2 |

| 1.9 In the last 6 months have you had any injury which was caused by work? | Yes **□** 1  No **□** 2 | **If yes,** please give details      **If no**, go to 1.10 | |
| --- | --- | --- | --- |
| **If yes,**  1.9a Was it reported to Workers Compensation? | | Yes **□** 1  No **□** 2 | |
| 1.9b Did it involve time off work, other than the day of the injury? | | Yes **□** 1  No **□** 2 | |
| 1.9c Did you receive any treatment for this? | | Yes **□** 1  No **□** 2 | **If yes**, was this treated by any of the following? (tick as many boxes as appropriate)  **□** On site first aid/occupational health  **□** Emergency department off site  **□** Walk-in clinic  **□** Family physician office  **□** Admitted to hospital  **□** Other, please give details    ____________________________ |

1.10 Do you have any other health problems now?	Yes	**□** 1	No **□** 2

**If no**, go to question 2

**If yes,** please give details.

| Nature of Problem | Was this made worse by work? | |
| --- | --- | --- |
| 1.10.1 | Yes | **□** 1 |
|  | No | **□** 2 |
| 1.10.2 | Yes | **□** 1 |
|  | No | **□** 2 |
| 1.10.3 | Yes | **□** 1 |
|  | No | **□** 2 |
| 1.10.4 | Yes | **□** 1 |
|  | No | **□** 2 |
| 1.10.5 | Yes | **□** 1 |
|  | No | **□** 2 |

2 **Are you taking any tablets or using other medication now (other than birth control)? Include both those prescribed by a doctor and those bought without prescription.**

Yes	**□** 1	No **□** 2

**If no,** go to question 3

**If yes,** please give details

| **What tablets or medication are you using?** | **How often do you take/use it?** | **For what condition or symptoms are you taking this medication?** |
| --- | --- | --- |
| 2.1 |  |  |
| 2.2 |  |  |
| 2.3 |  |  |
| 2.4 |  |  |

Reported harassment and mental-ill health in a Canadian prospective cohort of women and men in welding and electrical trades

Jean-Michel Galarneau, Quentin Durand-Moreau, Nicola Cherry

**Supplementary materials 2**

Long wrap-up questionnaire

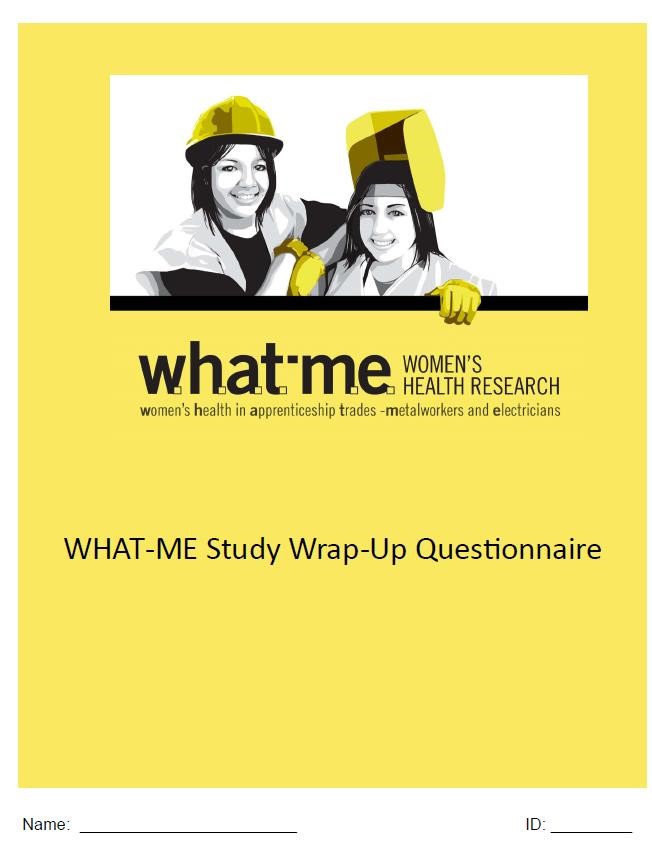

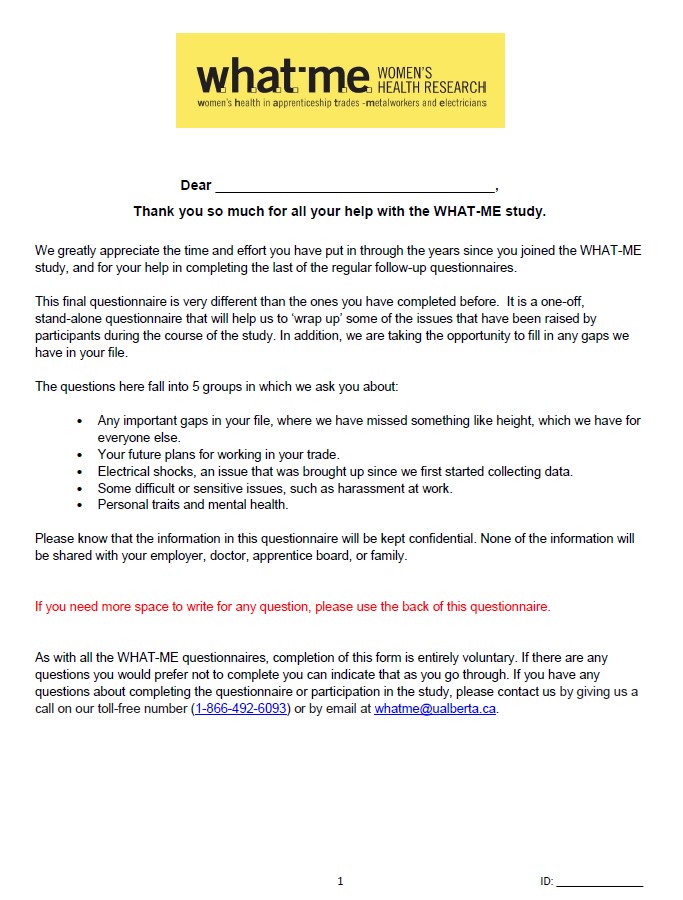

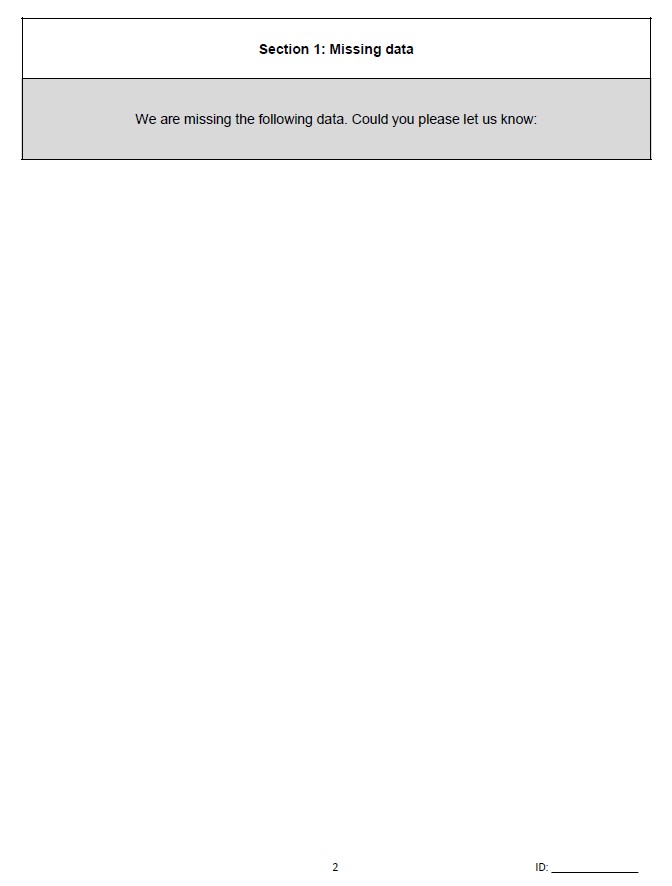

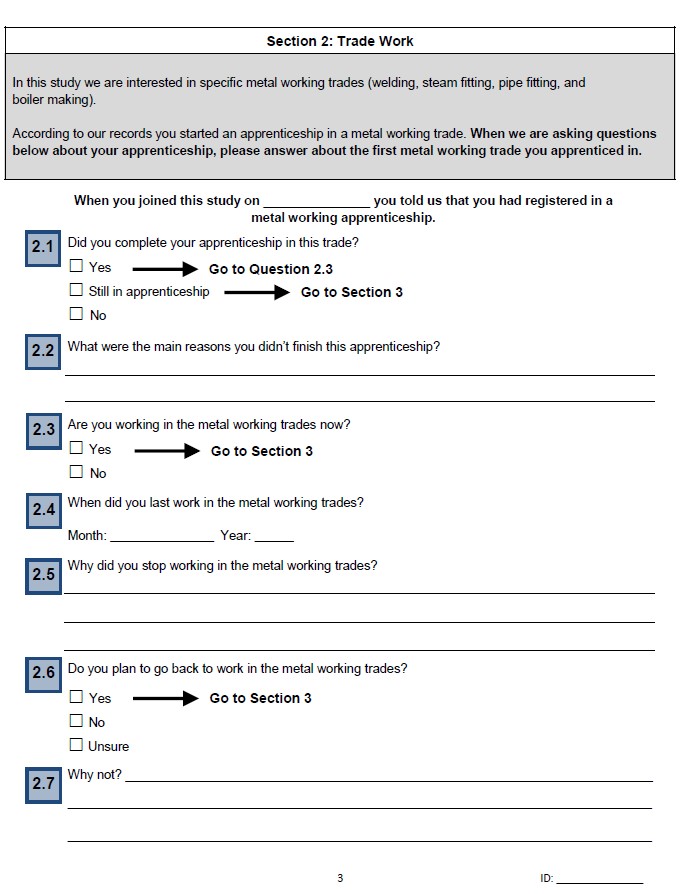

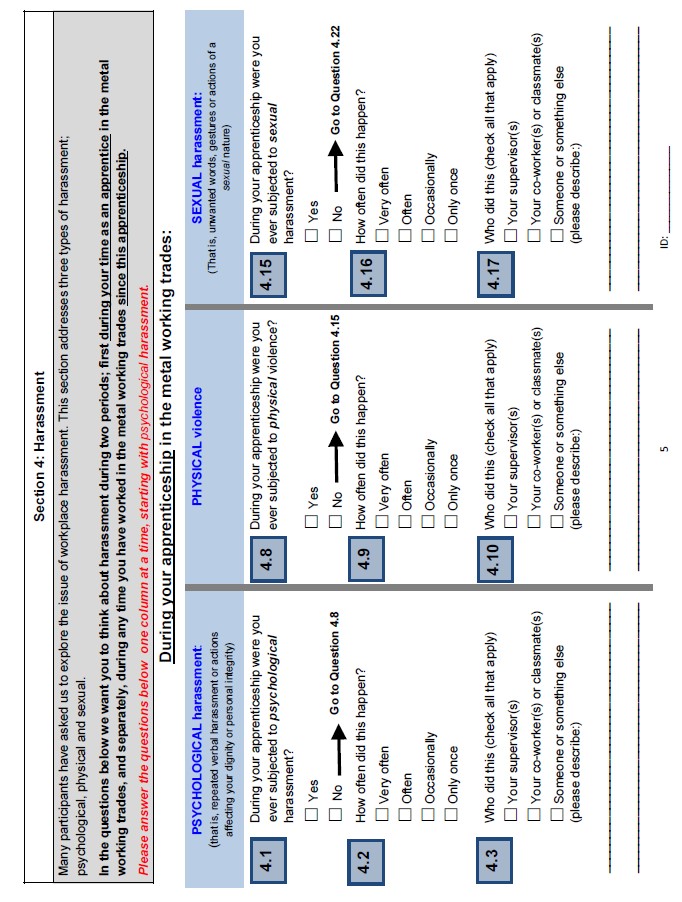

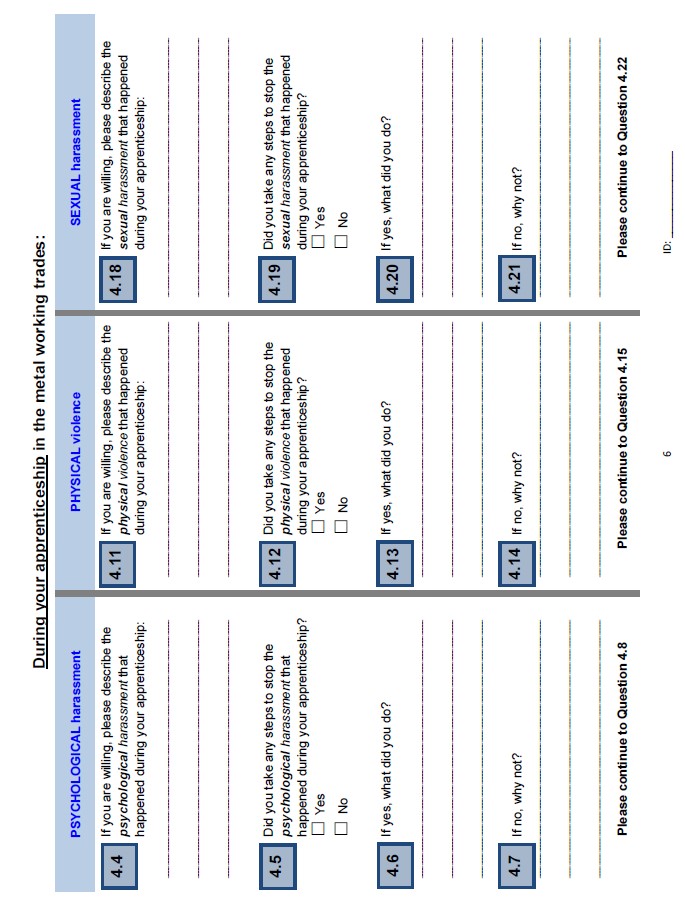

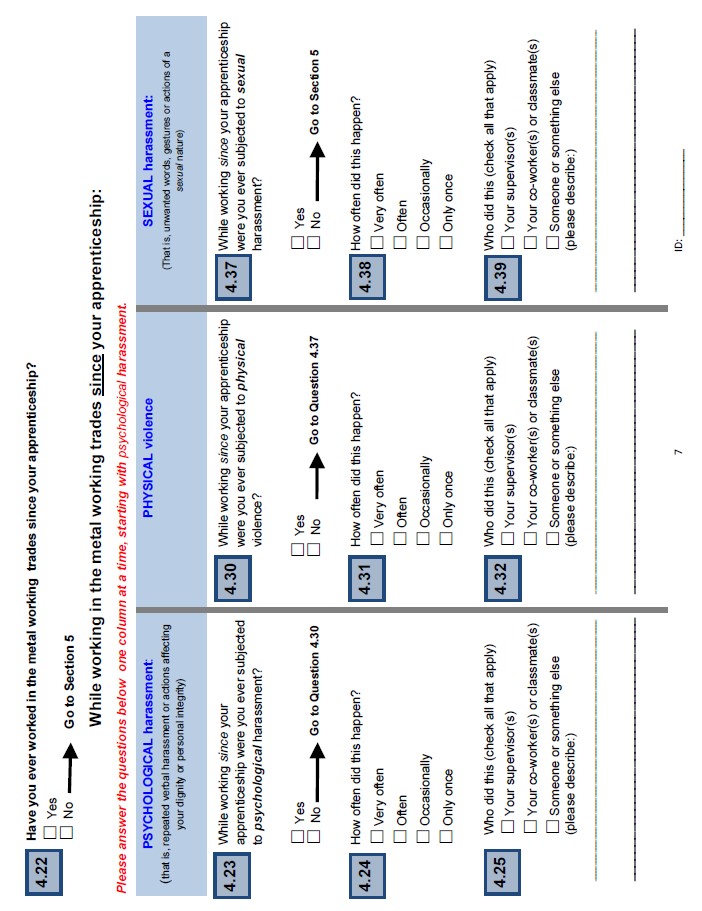

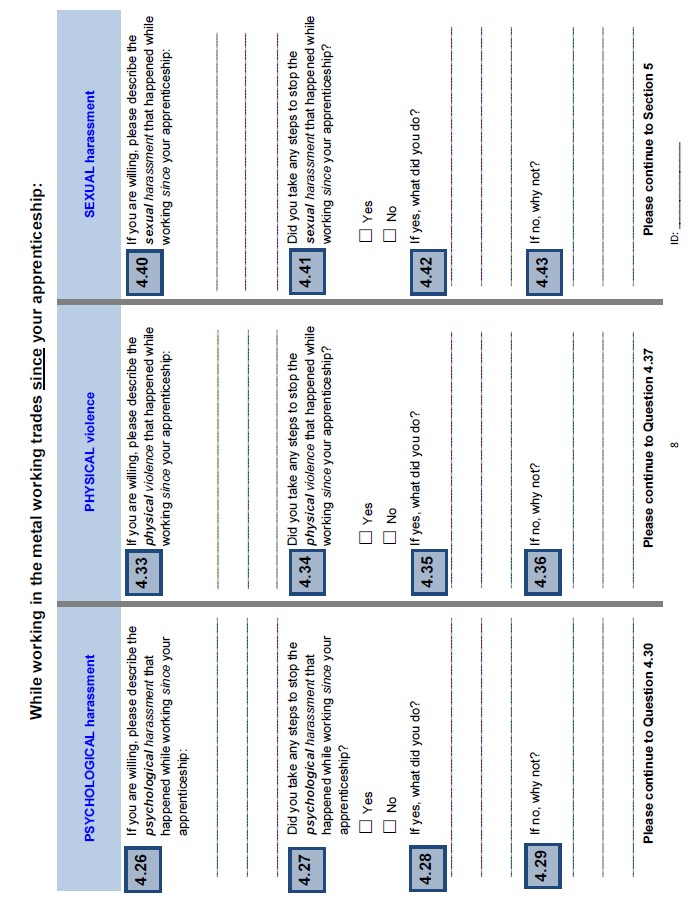

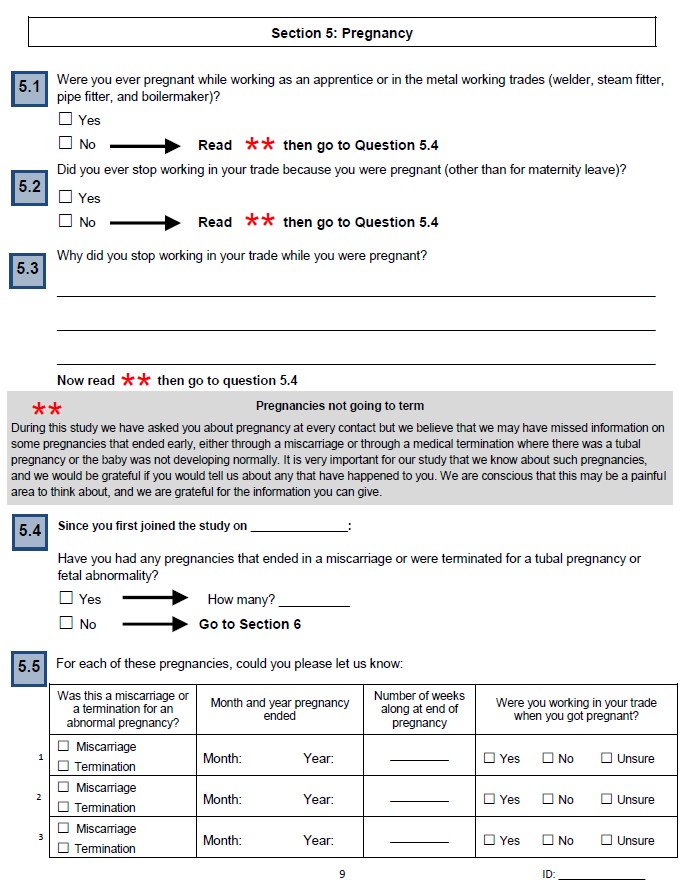

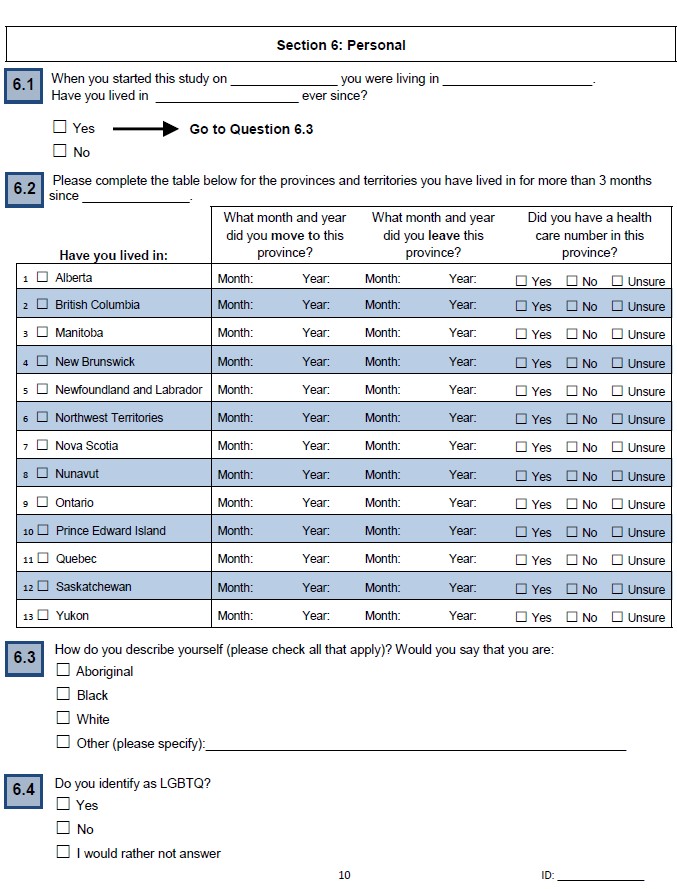

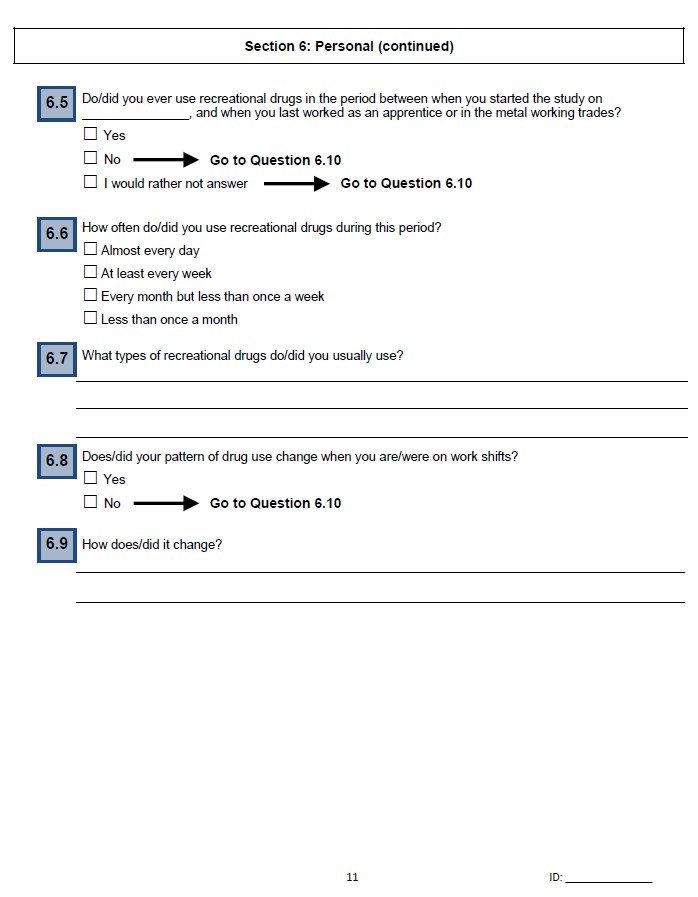

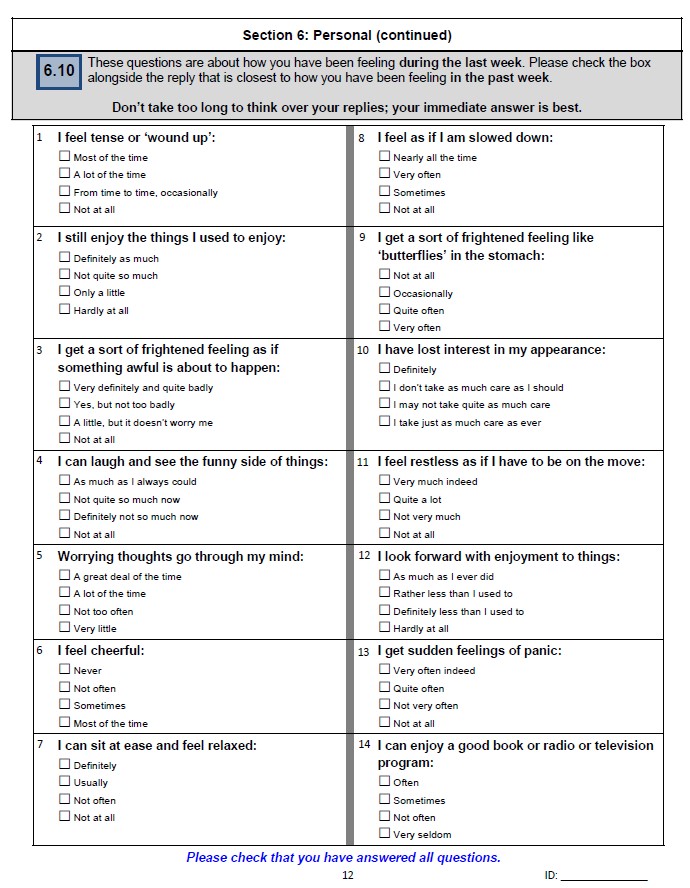

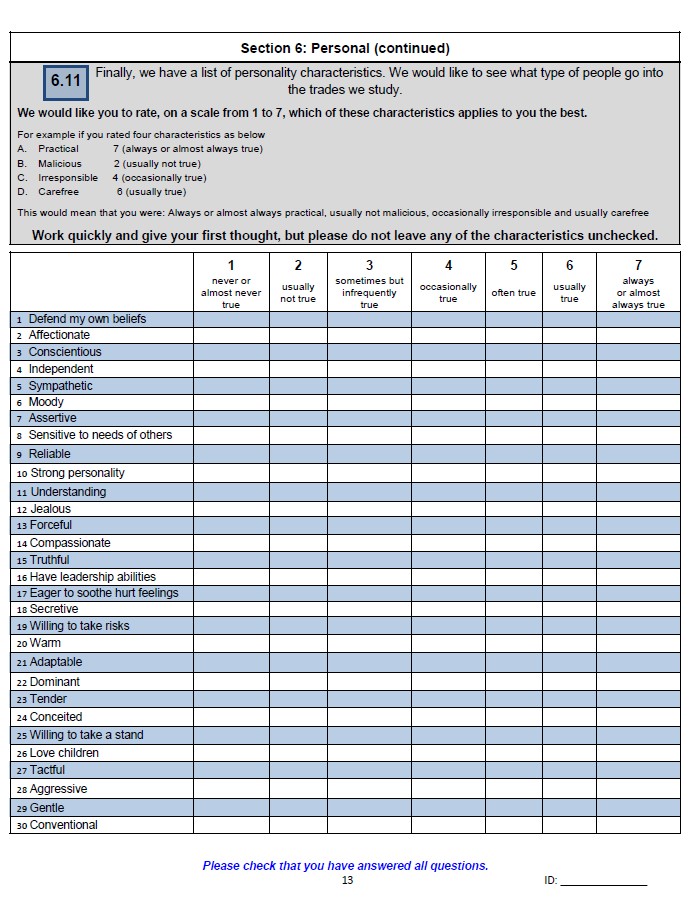

Reported harassment and mental-ill health in a Canadian prospective cohort of women and men in welding and electrical trades

Jean-Michel Galarneau, Quentin Durand-Moreau, Nicola Cherry

**Supplementary materials 3**

Short Wrap-up questionnaire

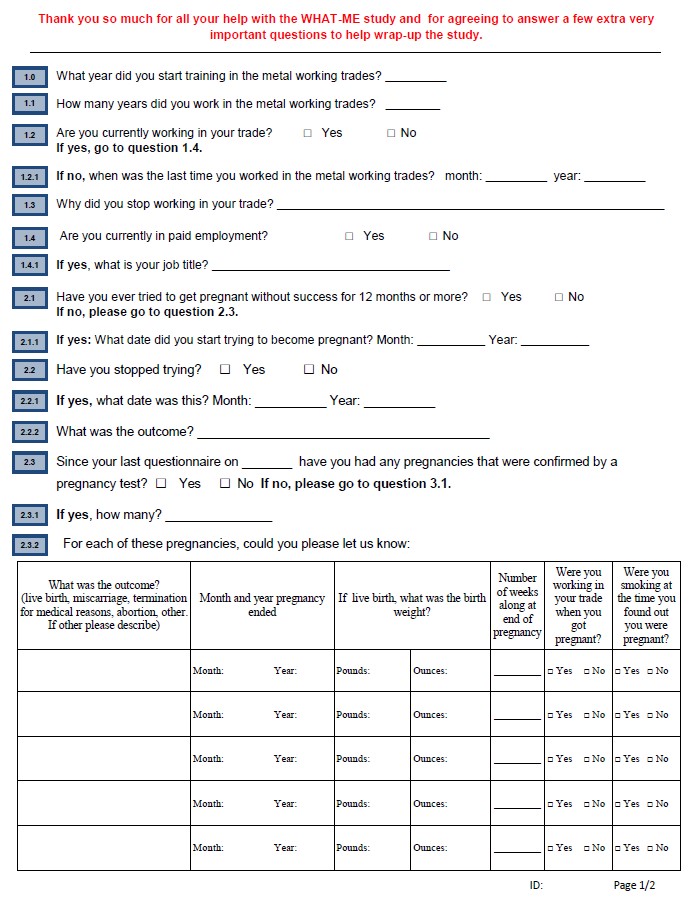

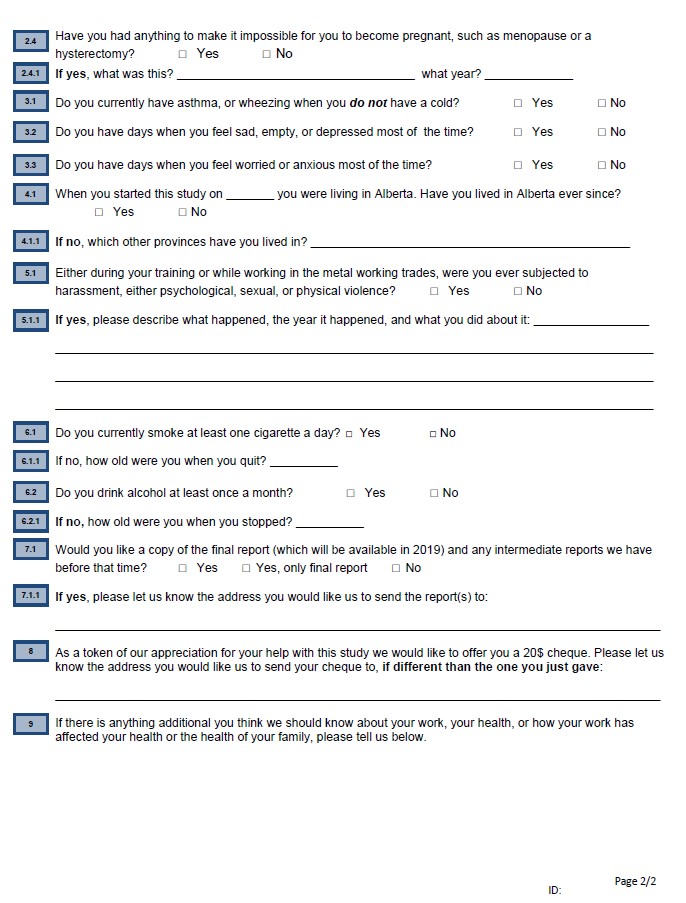

Reported harassment and mental-ill health in a Canadian prospective cohort of women and men in welding and electrical trades

Jean-Michel Galarneau, Quentin Durand-Moreau, Nicola Cherry

**Supplementary Materials 4**

Table SM1. Fractional regression of the proportion of periodic reports of anxiety or depression made worse by work in trade.

Table SM2. Relationship of reported harassment in trade to physician records of anxiety or depression before trade.

Table SM3. Multilevel logistic regression of periodic reports of anxiety and depression while in trade

Table SM4. Periodic reports of anxiety and depression with sexual harassment

Table SM5. Relationship of physician reported anxiety and depression to harassment since joining trade.

Table SM1. Fractional regression of the proportion of periodic reports of anxiety or depression made worse by work in trade.

|  | All (N=822) | | | Women (N=377) | | | Men (N=445) | | |
| --- | --- | --- | --- | --- | --- | --- | --- | --- | --- |
| Anxiety made worse by work | β | 95% CI | P= | β | 95% CI | P= | β | 95% CI | P= |
| Report of harassment | 0.37 | 0.21 to 0.53 | <0.001 | 0.37 | 0.12 to 0.62 | 0.003 | 0.37 | 0.17 to 0.58 | <0.001 |
| HADs anxiety score | 0.12 | 0.10 to 0.14 | <0.001 | 0.12 | 0.10 to 0.15 | <0.001 | 0.11 | 0.09 to 0.14 | <0.001 |
| Gender: man | 0.27 | 0.11 to 0.42 | 0.001 | - | - | - | - | - | - |
| Trade: welding | -0.06 | -0.21 to 0.09 | 0.432 | -0.14 | -0.38 to 0.10 | 0.257 | -0.00 | -0.20 to 0.19 | 0.978 |
| Total reports in trade | -0.03 | -0.07 to 0.01 | 0.132 | -0.06 | -0.12 to - 0.01 | 0.026 | -0.00 | -0.07 to 0.07 | 0.947 |
| Depression made worse by work |  |  |  |  |  |  |  |  |  |
| Report of harassment | 0.43 | 0.25 to 0.61 | <0.001 | 0.64 | 0.33 to 0.95 | <0.001 | 0.32 | 0.08 to 0.56 | 0.009 |
| HADs depression score | 0.09 | 0.75 to 0.12 | <0.001 | 0.08 | 0.05 to 0.12 | <0.001 | 0.10 | 0.07 to 0.13 | <0.001 |
| Gender: man | 0.15 | -0.03 to 0.32 | 0.112 | - | - | - | - | - | - |
| Trade: welding | 0.04 | -013 to 0.22 | 0.641 | 0.07 | -0.20 to 0.34 | 0.605 | 0.01 | -0.22 to 0.25 | 0.906 |
| Total reports in trade | -0.08 | -0.13 to -0.03 | 0.001 | -0.09 | -0.16 to -0.02 | 0.009 | -0.07 | -0.15 to 0.00 | 0.051 |

Table SM2. Relationship of reported harassment in trade to physician records of anxiety or depression before trade.

|  | Physician record of anxiety related condition before trade | | | | | | | | |
| --- | --- | --- | --- | --- | --- | --- | --- | --- | --- |
| Reported harassment | All | | | Women | | | Men | | |
|  | No | Yes | All | No | Yes | All | No | Yes | All |
| No | 476 | 76 | 552 | 119 | 33 | 152 | 357 | 43 | 400 |
| Yes | 308 | 86 | 394 | 149 | 56 | 205 | 159 | 30 | 189 |
| All | 784 | 162 | 946 | 268 | 89 | 357 | 516 | 73 | 589 |
| % | 39.3 | 53.1 | 41.6 | 55.6 | 62.9 | 57.4 | 30.8 | 41.1 | 32.1 |
| P | 0.002 | | | 0.266 | | | 0.083 | | |
|  | Physician record of depressive condition before trade | | | | | | | | |
| Reported harassment | All | | | Women | | | Men | | |
|  | No | Yes | All | No | Yes | All | No | Yes | All |
| No | 489 | 63 | 552 | 125 | 27 | 152 | 364 | 36 | 400 |
| Yes | 333 | 61 | 394 | 161 | 44 | 205 | 172 | 17 | 189 |
| All | 822 | 124 | 946 | 286 | 71 | 357 | 536 | 53 | 589 |
| % | 40.5 | 49.2 | 41.6 | 56.3 | 62.0 | 57.4 | 32.1 | 32.1 | 32.1 |
| P | 0.078 | | | 0.423 | | | 1.00 | | |

Table SM3. Multilevel logistic regression of periodic reports of anxiety and depression while in trade with anxiety or depression before trade from the administrative health database: Alberta sub-sample

|  | Periodic reports of anxiety or depression while in trade | | | | | | | | |
| --- | --- | --- | --- | --- | --- | --- | --- | --- | --- |
|  | All | | | Women | | | Men | | |
| Anxiety | OR | 95% CI | P= | OR | 95% CI | P= | OR | 95% CI | P= |
| Harassment | 1.97 | 1.28 to 3.04 | 0.002 | 1.53 | 0.76 to 3.06 | 0.230 | 2.09 | 1.21 to 3.61 | 0.008 |
| Anxiety before | 1.64 | 0.97 to 2.77 | 0.067 | 1.47 | 0.73 to 2.97 | 0.285 | 1.71 | 0.82 to 3.55 | 0.153 |
| HADS anxiety | 1.35 | 1.28 to 1.43 | <0.001 | 1.31 | 1.21 to 1.42 | <0.001 | 1.37 | 1.27 to 1.48 | <0.001 |
| Trade: welding | 0.85 | 0.56 to 1.28 | 0.427 | 0.66 | 0.35 to 1.25 | 0.203 | 0.96 | 0.56 to 1.65 | 0.889 |
| Sex: male | 1.89 | 1.20 to 2.98 | 0.006 | - | - | - | - | - | - |
| Depression | | | | | | | | | |
| Harassment | 2.09 | 1.24 to 3.53 | 0.006 | 3.09 | 0.98 to 9.69 | 0.054 | 1.95 | 1.05 to 3.61 | 0.034 |
| Depression before | 0.94 | 0.48 to 1.84 | 0.847 | 1.87 | 0.57 to 6.06 | 0.299 | 0.72 | 0.31 to 1.68 | 0.451 |
| HADS depression | 1.37 | 1.28 to 1.47 | <0.001 | 1.22 | 1.05 to 1.41 | 0.010 | 1.39 | 1.28 to 1.50 | <0.001 |
| Trade: welding | 1.05 | 0.65 to 1.69 | 0.847 | 1.71 | 0.74 to 3.95 | 0.206 | 1.02 | 0.55 to 1.88 | 0.960 |
| Sex: male | 1.00 | 0.61 to 1.64 | 0.990 | - | - | - | - | - | - |
| N observations | 2831 | - | - | 839 | - | - | 1992 | - | - |
| Participants | 577 | - | - | 198 | - | - | 379 | - | - |
|  | Periodic reports of anxiety or depression made worse by work (in those reporting symptoms) | | | | | | | | |
|  | All | | | Women | | | Men | | |
| Anxiety | OR | 95% CI | P= | OR | 95% CI | P= | OR | 95% CI | P= |
| Harassment | 2.18 | 1.24 to 3.83 | 0.007 | 3.72 | 1.21 to 11.41 | 0.022 | 1.81 | 0.94 to 3.48 | 0.077 |
| Anxiety before | 0.66 | 0.35 to 1.27 | 0.216 | 0.49 | 0.18 to 1.38 | 0.177 | 0.77 | 0.32 to 1.81 | 0.545 |
| HADS anxiety | 1.15 | 1.07 to 1.24 | <0.001 | 1.24 | 1.09 to 1.42 | 0.001 | 1.10 | 1.01 to 1.21 | 0.036 |
| Trade: welding | 0.69 | 0.40 to 1.19 | 0.183 | 0.61 | 0.23 to 1.63 | 0.322 | 0.71 | 0.37 to 1.35 | 0.294 |
| Sex: male | 2.62 | 1.42 to 4.84 | 0.002 | - | - | - | - | - | - |
| N observations | 656 | - | - | 195 | - | - | 461 | - | - |
| Participants | 280 | - | - | 93 | - | - | 187 | - | - |
| Depression | | | | | | | | | |
| Harassment | 2.10 | 1.10 to 3.99 | 0.024 | 7.33 | 2.24 to 24.01 | 0.001 | 1.37 | 0.64 to 2.91 | 0.419 |
| Depression before | 0.35 | 0.14 to 0.89 | 0.027 | 0.29 | 0.09 to 0.93 | 0.037 | 0.35 | 0.09 to 1.40 | 0.139 |
| HADS depression | 1.07 | 0.99 to 1.16 | 0.085 | 1.04 | 0.90 to 1.20 | 0.592 | 1.08 | 0.98 to 1.19 | 0.103 |
| Trade: welding | 0.89 | 0.48 to 1.63 | 0.696 | 1.43 | 0.56 to 3.65 | 0.455 | 0.75 | 0.35 to 1.59 | 0.454 |
| Sex: male | 1.27 | 0.63 to 2.55 | 0.509 | - | - | - | - | - | - |
| N observations | 510 | - | - | 158 | - | - | 352 | - | - |
| Participants | 229 | - | - | 75 | - | - | 154 | - | - |

Table SM4. Periodic reports of anxiety and depression with sexual harassment (Logistic regression)

|  | Periodic reports of anxiety or depression while in trade | | | | | | | | |
| --- | --- | --- | --- | --- | --- | --- | --- | --- | --- |
|  | All | | | Women | | | Men | | |
| Anxiety | OR | 95% CI | P= | OR | 95% CI | P= | OR | 95% CI | P= |
| Harassment |  |  |  |  |  |  |  |  |  |
| None | 1.00 |  |  | 1.00 |  |  | 1.00 |  |  |
| Sexual: no | 1.76 | 1.13 to 2.72 | 0.012 | 0.95 | 0.45 to 2.00 | 0.890 | 2.22 | 1.31 to 3.73 | 0.003 |
| Sexual: yes | 1.98 | 1.19 to 3.28 | 0.009 | 1.27 | 0.71 to 2.29 | 0.422 | 6.77 | 1.91 to 23.99 | 0.003 |
| HADS anxiety | 1.37 | 1.30 to 1.44 | <0.001 | 1.35 | 1.26 to 1.44 | <0.001 | 1.40 | 1.30 to 1.50 | <0.001 |
| Trade: welding | 0.95 | 0.67 to 1.36 | 0.780 | 0.88 | 0.53 to 1.46 | 0.625 | 1.03 | 0.62 to 1.71 | 0.899 |
| Gender: man | 1.60 | 1.05 to 2.45 | 0.030 | - | - | - | - | - | - |
| Depression | | | | | | | | | |
| Harassment |  |  |  |  |  |  |  |  |  |
| None | 1.00 |  |  | 1.00 |  |  | 1.00 |  |  |
| Sexual: no | 2.77 | 1.64 to 4.68 | <0.001 | 5.65 | 2.44 to 13.11 | <0.001 | 1.79 | 0.96 to 3.36 | 0.068 |
| Sexual: yes | 2.85 | 1.59 to 5.11 | <0.001 | 3.61 | 1.63 to 8.02 | 0.002 | 6.64 | 2.38 to 18.51 | <0.001 |
| HADS depression | 1.37 | 1.29 to 1.46 | <0.001 | 1.42 | 1.29 to 1.57 | <0.001 | 1.41 | 1.30 to 1.52 | <0.001 |
| Trade: welding | 1.43 | 0.95 to 2.15 | 0.086 | 1.62 | 0.92 to 2.82 | 0.092 | 1.45 | 0.82 to 2.57 | 0.203 |
| Gender: man | 1.07 | 0.68 to 1.69 | 0.775 | - | - | - | - | - | - |
| N observations | 4022 | - | - | 1718 | - | - | 2304 | - | - |
| Participants | 821 | - | - | 377 | - | - | 444 | - | - |
|  | Periodic reports of anxiety or depression made worse by work (in those reporting symptoms) | | | | | | | | |
|  | All | | | Women | | | Men | | |
| Anxiety | OR | 95% CI | P= | OR | 95% CI | P= | OR | 95% CI | P= |
| Harassment |  |  |  |  |  |  |  |  |  |
| None | 1.00 | - | - | 1.00 | - | - | 1.00 | - | - |
| Sexual: no | 2.07 | 1.20 to 3.56 | 0.009 | 4.02 | 1.51 to 10.71 | 0.005 | 1.57 | 0.80 to 3.06 | 0.187 |
| Sexual: yes | 2.04 | 1.09 to 3.83 | 0.026 | 2.27 | 1.07 to 4.83 | 0.033 | 2.61 | 0.67 to 10.25 | 0.168 |
| HADS anxiety | 1.14 | 1.07 to 1.21 | <0.001 | 1.16 | 1.07 to 1.26 | 0.001 | 1.11 | 1.01 to 1.22 | 0.027 |
| Trade: welding | 0.69 | 0.43 to 1.09 | 0.109 | 0.67 | 0.35 to 1.29 | 0.236 | 0.71 | 0.37 to 1.35 | 0.293 |
| Gender: man | 2.02 | 1.18 to 3.44 | 0.010 | - | - | - | - | - | - |
| N observations | 942 | - | - | 420 | - | - | 522 | - | - |
| Participants | 396 | - | - | 184 | - | - | 212 | - | - |
| Depression | | | | | | | | | |
| Harassment |  |  |  |  |  |  |  |  |  |
| None | 1.00 | - | - | 1.00 | - | - | 1.00 | - | - |
| Sexual: no | 1.91 | 1.04 to 3.51 | 0.036 | 3.41 | 1.23 to 9.44 | 0.018 | 1.56 | 0.72 to 3.38 | 0.257 |
| Sexual: yes | 3.76 | 1.74 to 8.10 | 0.001 | 5.98 | 2.35 to 15.20 | <0.001 | 1.64 | 0.35 to 7.68 | 0.533 |
| HADS depression | 1.09 | 1.02 to 1.17 | 0.017 | 1.10 | 0.98 to 1.22 | 0.107 | 1.09 | 0.99 to 1.19 | 0.079 |
| Trade: welding | 1.03 | 0.62 to 1.74 | 0.904 | 1.26 | 0.62 to 2.57 | 0.521 | 0.87 | 0.42 to 1.80 | 0.710 |
| Gender: man | 1.86 | 1.01 to 3.43 | 0.047 | - | - | - | - | - | - |
| N observations | 722 | - | - | 312 | - | - | 410 | - | - |
| Participants | 323 | - | - | 143 | - | - | 180 | - | - |

Table SM5. Relationship of physician reported anxiety and depression to harassment since joining trade.

|  | Reported harassment | | | | | | | | |
| --- | --- | --- | --- | --- | --- | --- | --- | --- | --- |
| Physician recorded | All | | | Women | | | Men | | |
|  | No | Yes | All | No | Yes | All | No | Yes | All |
| Anxiety |  |  |  |  |  |  |  |  |  |
| No | 311 | 185 | 496 | 72 | 88 | 160 | 239 | 97 | 336 |
| Yes | 241 | 209 | 450 | 80 | 117 | 197 | 161 | 92 | 253 |
| All | 552 | 394 | 946 | 152 | 205 | 357 | 400 | 189 | 589 |
| % | 43.7 | 53.0 | 47.6 | 52.6 | 57.1 | 55.2 | 40.3 | 48.7 | 43.0 |
| P | 0.005 | | | 0.451 | | | 0.061 | | |
| Depression |  |  |  |  |  |  |  |  |  |
| No | 430 | 241 | 671 | 112 | 119 | 231 | 318 | 122 | 440 |
| Yes | 122 | 153 | 275 | 40 | 86 | 126 | 82 | 67 | 149 |
| All | 552 | 394 | 946 | 152 | 205 | 357 | 400 | 189 | 589 |
| % | 22.1 | 38.8 | 29.1 | 26.3 | 42.0 | 35.3 | 20.5 | 35.4 | 25.3 |
| P | 0.001 | | | 0.002 | | | <0.001 | | |

Reported harassment and mental-ill health in a Canadian prospective cohort of women and men in welding and electrical trades

Jean-Michel Galarneau, Quentin Durand-Moreau, Nicola Cherry

**Supplementary materials 5:** Calculation of misreporting needed to eliminate a result.

1) A total of 1413 completed the harassment questionnaire of whom 1187 (84%) completed at least one periodic report about anxiety or depression made worse by work while they were working in their trade
2) 707 of those reporting while in trade never reported symptoms of anxiety or depression made worse by work in follow-up from 2011. Among this group 36% (253/707) reported harassment in 2016-18
3) 480 of those reporting while in trade reported either anxiety or depression made worse by work on at least one occasion at follow-up from 2011-2018. Among this group 278 (58%) reported harassment in 2016-2018.
4) From these data the unadjusted odds ratio for anxiety or depression made worse by work was 2.47 95%CI 1.95 to 3.13. Adjusted for sex, HADS anxiety and depression, trade and total number of reports the OR reduces slightly to 2.23 95%CI 1.59-3.13 (for the sub-group with HADS scores, N=822).
5) To reduce the unadjusted OR to 1.00 with 95%CI: 0.79 to 1.28 (that is no effect of harassment on anxiety or depression made worse by work) 106 of 278 (37%) reporting anxiety or depression would have had to be a false positive on harassment (leaving 174/480 36.5% as true cases), assuming little or no error in the reporting of harassment in those who never reported anxiety or depression made worse by work.
